# Subclinical hepatorenal biomarker alterations and urogenital indicators in *Schistosoma haematobium*-infected School-Aged Children in Sene West District, Ghana

**DOI:** 10.64898/2026.09.30.26364430

**Authors:** Dennis Kyei Ofori, Anabel Acheampong, Prince-Charles Kudzordzi, Prince Nyarko, Claudia Wubuareyasa Nseide, Emmanuel Ansah Boateng, Yahuza Sabit Tanko, Conor R. Caffrey, Kenneth Bentum Otabil

## Abstract

Urogenital schistosomiasis, caused by *Schistosoma haematobium*, remains a major public health challenge among school-aged children in rural Ghana. Although urogenital indicators are well-established, systemic hepatic biomarker alterations in post-mass drug administration pediatric settings remain poorly characterized. This cross-sectional study evaluated point-of-care urinalysis profiles alongside a panel of serum hepatic and renal biomarkers in 174 children (aged 3–16 years) from the Sene West District, Ghana. Active infection (urine microscopy egg detection) was identified in 84 participants (48.3%, median egg count = 4.50 eggs/10 mL, IQR: 1.00–9.25). Active infection was strongly associated with microhematuria (57.1% vs. 11.1%, p < 0.001), proteinuria (42.9% vs. 22.2%, *p* = 0.004), and leukocyturia (27.4% vs. 4.4%, *p* < 0.001). Children with active infection also demonstrated a higher frequency of composite categorical hepatic biomarker alterations (60.7% vs. 37.8%, *p* = 0.002), whereas categorical renal alterations showed no significant difference (*p* = 0.332). Controlling for age and sex, multivariable logistic regression confirmed active infection as a significant independent predictor of composite hepatic biomarker alterations (aOR = 2.54, 95% CI: 1.37–4.69, p = 0.003), but not renal alterations (aOR = 1.40, p = 0.359). Continuous medians for individual serum biomarkers showed no significant differences between groups. Combining composite hepatic biomarker thresholds with routine dipstick screening improves the detection of subclinical morbidity in pediatric field settings.

## Background

Schistosomiasis is one of the most socioeconomically destructive parasitic diseases globally, second only to malaria, and accounts for over 200,000 deaths annually (Global Burden of Disease Injury Prevention and Control Collaborators, 2018). Worldwide, over 240 million people require preventive chemotherapy for schistosomiasis, with nearly 90% of cases concentrated in Sub-Saharan Africa (World Health Organization, 2020). Urogenital schistosomiasis, caused by the blood fluke *Schistosoma haematobium,* accounts for over half of the global schistosomiasis burden and remains a major public health problem across the African continent (WHO, 2020; Vere et al., 2025). The disease has been earmarked for elimination as a public health problem (EPHP) by 2030 under the World Health Organization’s (WHO) neglected tropical disease roadmap (WHO, 2020).

Transmission of *S. haematobium* is driven by human contact with infested freshwater bodies containing infective larvae (cercariae) that are shed by intermediate snail hosts (*Bulinus* spp.) (WHO, 2020). Following skin penetration and subsequent intravascular migration, adult worm pairs colonize the vesical and pelvic venous plexuses. Female worms release hundreds of eggs daily into surrounding tissues, provoking intense cell-mediated granulomatous inflammation and progressive collagen deposition (Shwartz and Fallon, 2018) Locally, this tissue injury manifests as microhematuria, proteinuria, leukocyturia, dysuria, and pollakiuria (Vere et al., 2025). Beyond localized urogenital lesions, chronic inflammatory signaling, circulating immune complexes, and altered hemodynamic responses can induce secondary biomarker alterations in distal organ systems, including the liver and kidneys (Gryseels et al, 2006; Manciulli et al, 2023; WHO, 2026; Bernado et all., 2020).

Routine serum biochemistry panels measuring systemic hepatorenal biomarkers (*e.g*., transaminases, serum creatinine, urea, and electrolytes) can detect early infection-associated biomarker alterations before overt structural organ damage ensues (Manciulli et al., 2023). However, schistosomiasis control programmes do not incorporate systemic laboratory testing due to resource constraints (Rollinson et al., 2013). Instead, field surveillance and mass drug administration campaigns rely almost exclusively on microscopic egg quantification or isolated urinalysis dipstick parameters (Rollinson et al., 2013). Point-in-time egg counts, or single binary dipstick indicators may fail to reflect cumulative tissue responses because a substantial proportion of eggs remain trapped in host tissues rather than excreted in urine. Consequently, a diagnostic reliance on localized excretory markers potentially masks underlying subclinical biomarker alterations in endemic pediatric populations. This would underscore the clinical value of evaluating systemic hepatorenal profiles alongside routine dipstick screening.

The existing literature evaluating systemic organ involvement in schistosomiasis-endemic communities reports conflicting findings (Abdelghani et al., 2020; Legido-Quigley, 2010). These discrepancies span both hepatic (ALT, AST, ALP, gamma-GGT, bilirubin) and renal markers (serum creatinine, urea, eGFR) (Abdelghani et al., 2020; Legido-Quigley, 2010). Driven by varying parasite burdens, exposure durations, host demographics, and underlying co-infections, findings from one epidemiological setting cannot be generalized to another. Consequently, subclinical hepatorenal alterations frequently go undetected in high-burden communities (Abdelghani et al., 2020; Legido-Quigley, 2010). Rigorous field-based evaluations that integrate local urogenital pathology metrics (specifically urine egg presence, microhematuria, proteinuria, and leukocyturia) with systemic hepatorenal biomarker profiling are, therefore, essential to better characterize the true multi-system impact of pediatric schistosomiasis.

This study aimed to evaluate local urogenital dipstick parameters, systemic hepatic and renal biomarker profiles, as well as the overall patterns of their alterations among school-aged children in an *S. haematobium*-endemic area, namely the Sene West District of Ghana.

## Materials and methods

### Study area and setting

The study was conducted across four urinary schistosomiasis-endemic rural communities: Tato Battor, Kpatsa, Gobu, and Kwaese located in the Sene West District of the Bono East Region, Ghana (Figure 1). These communities are situated along freshwater bodies contiguous with Volta Lake and characterized by limited sanitation and hygiene (WASH) infrastructure (Grimes et al., 2015), necessitating heavy reliance on surface water for domestic, recreational, and occupational activities. The primary livelihoods, including subsistence farming, artisanal fishing and sand- winning, involve frequent, and often prolonged, contact with natural water bodies that harbour the *Bulinus* intermediate snail host of *S. haematobium*. The interplay between high human-water contact, inadequate WASH services, and favorable ecological conditions for vector snails sustains active transmission (Grimes et al., 2015), providing an ideal epidemiological setting to evaluate *S. haematobium*-associated hepatorenal biomarker alterations.

**Figure 1.**
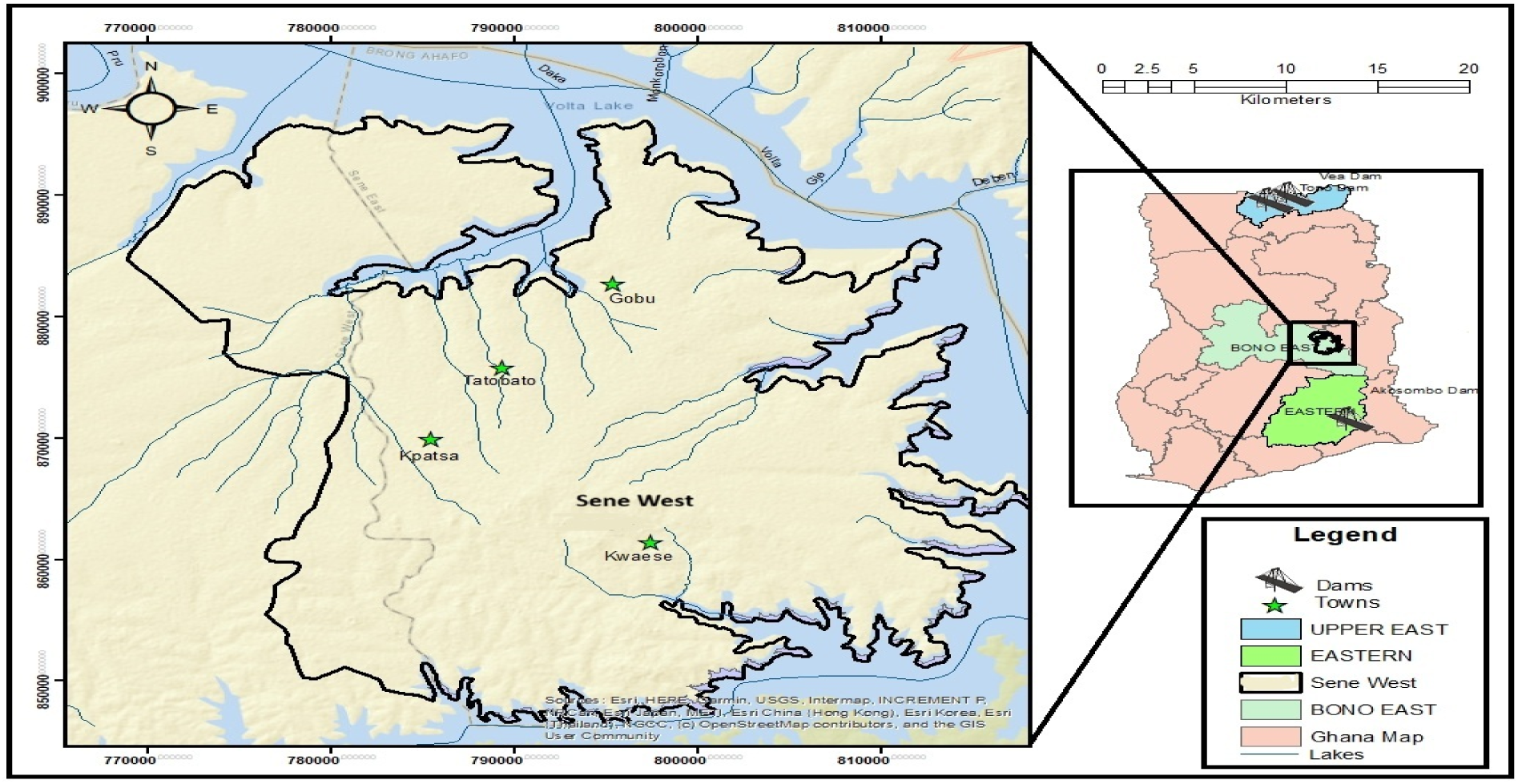
Map of Ghana showing the study areas in the Sene West District of the Bono East Region of Ghana.

### Study design

A community-based cross-sectional study was conducted among children and adolescents aged 3–16 years residing in the four endemic communities (Table 1). Infection status (microscopic egg presence), point-of-care urinalysis parameters (microhematuria, proteinuria, and leukocyturia), serum hepatorenal biomarkers, and electrolytes were evaluated concurrently in all enrolled participants. This design enabled the determination of *S. haematobium* prevalence while simultaneously assessing infection-associated differences in localized urogenital indicators and systemic biomarker profiles across the cohort.

**Table 1.** Demographic and community baseline characteristics of the study population.

| Characteristic / Variable | Frequency (n) | Percentage (%) |
| --- | --- | --- |
| **Sex** |  |  |
| Female | 85 | 48.9% |
| Male | 89 | 51.1% |
| **Age stratification (WHO/pediatric brackets)** |  |  |
| 3–4 years (Preschool / Early Childhood) | 16 | 9.2% |
| 5–9 years (Early Primary School) | 88 | 50.6% |
| 10–12 years (Late Primary / Early Adolescence) | 33 | 19.0% |
| 13–16 years (Junior High / Adolescence) | 37 | 21.3% |
| **Study site / community** |  |  |
| Tato Battor | 87 | 50.0% |
| Kpatsa | 39 | 22.4% |
| Gobu | 35 | 20.1% |
| Kwaese | 13 | 7.5% |

*Note. Relative proportions calculated on Total N = 174. Age range: 3–16 years.*

### Ethical clearance

The study protocol was reviewed and approved by the Committee for Human Research Ethics (CHRE) at the University of Energy and Natural Resources, Sunyani, Ghana (CHRE/CA/025/022).

### Inclusion and exclusion criteria

We enrolled children and adolescents aged 3–16 years from primary and junior high schools across four disease-endemic communities: Tato Battor, Kpatsa, Gobu, and Kwaese (Figure 1) (Opare et al., 2025). Participation required written informed consent from parents or legal guardians, and child assent for individuals aged 10–16 years. All enrolled participants provided paired urine samples (for parasitological egg counts and dipstick chemistry) and venous blood samples (for hepatorenal biomarker analysis). We excluded individuals with self- or parent/guardian-reported histories of pre-existing, non-schistosomal chronic liver or kidney disease (such as viral hepatitis), known congenital urinary tract anomalies, or praziquantel (PZQ) administration within the preceding 4–6 weeks. Participants with incomplete laboratory or demographic records were omitted from the final analysis.

### Sampling technique and sample size determination

Participants were recruited using a multi-stage probability sampling design across four rural communities in the Sene West District (Figure 1). Community selection was informed by preliminary district epidemiological surveys conducted in May 2025, which confirmed high baseline *S. haematobium* endemicity, with prevalence ranging between 40.0 and 79.0% (Acheampong et al., unpublished data). In November 2025, a school-based mass drug administration (MDA) campaign using PZQ was implemented district-wide by the Ghana Health Service. Sampling for the present study was conducted in February 2026, three months post-MDA. Within primary and junior high schools in each community, official class rosters served as the sampling frame.

To verify statistical power, sample size adequacy was evaluated *a priori* using Cochran’s formula (Daniel & Cross, 2018) for observational cross-sectional studies:

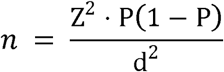

where Z = 1.96 (corresponding to a 95% confidence level), P = 0.50 (assumed baseline proportion to yield maximum sample size), and margin of error d = 0.075 (7.5%). This yielded a minimum target sample size of n = 171. The final systematically selected sample of N = 174 pupils (n = 90 microscopy negative, n = 84 microscopy positive) provided sufficient statistical power (≥ 80%) to detect cross-sectional differences in local urogenital dipstick indicators, serum biomarker alterations, and multivariable logistic regression odds ratios.

### Urine collection, dipstick urinalysis, and parasitology

Field sampling was conducted on-site across participating schools. Approximately 10 mL of mid-day urine was collected from each child between 10:00 AM and 2:00 PM to coincide with peak diurnal *S. haematobium* egg excretion (WHO, 1991). Under the supervision of trained research assistants, children were instructed to collect the terminal stream—the final portion of micturition—to maximize egg recovery (WHO, 1991). Point-of-care urinalysis was performed immediately using the Mission 10-in-1 reagent strips (Acon Labs, USA) according to the manufacturer’s protocol. Dipstick parameters (microhematuria, proteinuria, and leukocyturia) were graded visually using standardized colorimetric thresholds prior to microscopic egg quantification.

Urine specimens were immediately placed in a vehicle-powered mobile refrigerator (4–8 °C) to arrest egg hatching during transport from the school collection sites to the laboratory (a transit time of approximately 4 h). All specimens were processed on the day of collection or within 24 h, in accordance with standard diagnostic protocols (WHO, 1991). *S. haematobium* eggs were detected and quantified via urine sedimentation microscopy (WHO, 1991). Briefly, 10 mL of thoroughly homogenized urine was centrifuged at 2,000 × g for 2 min. Following careful decanting of the supernatant, the resuspended sediment was transferred to clean glass slides and systematically examined under 10× and 40× light microscopy to quantify terminal-spined *S. haematobium* eggs. Infection intensity was expressed as eggs per 10 mL of urine (eggs/10 mL) and classified according to WHO criteria: light infection (< 50 eggs/10 mL) and heavy infection (≥ 50 eggs/10 mL) (WHO, 2022).

### Blood collection and laboratory biochemical assays

Following urine collection, approximately 5 mL of venous blood was drawn aseptically from the antecubital fossa of each participant by a certified phlebotomist using sterile equipment. Samples were collected into serum separator tubes (SST), allowed to clot undisturbed at room temperature for 30 min, and centrifuged at 2,000 × g for 10 min. The resulting serum was carefully aliquoted into sterile cryovials and stored at -20 °C pending biochemical analysis.

Serum hepatic (ALT, AST, ALP, γ-GGT, total bilirubin) and renal biomarkers (creatinine, urea, eGFR), and electrolytes (Na, K, Cl) were quantified using a DS-161 Auto Biochemistry Analyzer (SINNOWA Medical Science & Technology Co., Ltd., Nanjing, China) according to the manufacturer’s protocol. System calibration was performed prior to sample processing, and commercial internal quality control (IQC) materials across normal and pathological ranges were analyzed daily to ensure analytical accuracy and precision. All laboratory operations adhered to standard operating procedures and Good Clinical Laboratory Practice (GCLP) guidelines (Ezzelle et al., 2007).

### Operational definitions and biomarker stratification

Organ-specific composite panel (more than one biomarker) alterations were established using pediatric age-adjusted clinical reference cutoffs:

- **Composite hepatic biomarker alterations:** Defined as a binary outcome (0 = No, 1 = Yes) indicating an elevation above age-adjusted upper reference limits in any key liver enzyme (ALT, AST, γ-GGT, or ALP) or total bilirubin, based on standard pediatric reference thresholds (Rifai, 2017).
- **Composite renal biomarker alterations:** Defined as a binary outcome (0 = No, 1 = Yes) based on serum creatinine > 100 µmol/L, elevated urea, or eGFR < 60 mL/min/1.73 m², referencing KDIGO (Kidney Disease: Improving Global Outcomes) pediatric guidance (KDIGO, 2013).
- **Estimated Glomerular Filtration Rate (eGFR):** Calculated using the Bedside Schwartz equation for pediatric participants (Schwartz et al., 2009).
- **De Ritis ratio:** this is calculated as the ratio of serum AST to ALT, and evaluated as both a continuous variable and a binary indicator (< 1.0 vs. ≥ 1.0) (Botros and Sikaris, 2013). This ratio serves as an indicator of the depth of cellular liver injury, where values < 1.0 reflect mild cytoplasmic leakage and values ≥ 1.0 indicate deeper mitochondrial damage or broader systemic involvement.

### Statistical analysis

Data were entered using Microsoft Excel (Microsoft 365) and analyzed using Jamovi (Version 2.7). Continuous variables were evaluated for distributional normality using the Shapiro-Wilk test. Because continuous biomarker distributions deviated significantly from normality (p < 0.05), they were summarized as medians and interquartile ranges (IQR: Q1–Q3). Between-group continuous comparisons across *S. haematobium* infection status (n = 84 microscopy-positive vs. n = 90 microscopy-negative) were evaluated using the non-parametric Mann-Whitney U test.

Categorical variables (including demographic characteristics, point-of-care urinalysis parameters, and binary biomarker alterations) were summarized as absolute counts (n) and percentages (%). Differences in categorical proportions by infection status were analyzed using Pearson’s Chi-square (χ²) test, or Fisher’s exact test when expected cell counts were fewer than five (5).

Multivariable binary logistic regression models were constructed to evaluate independent associations while adjusting for potential demographic confounders (age and sex). Binary logistic regression examined independent associations between *S. haematobium* infection status (positive vs. negative), sex (male vs. female), and age (years) with specific binary composite biomarker outcomes [Model 4A: Composite hepatic biomarker alterations (Yes vs. No); Model 4B: Composite renal biomarker alterations (Yes vs. No); Model 4C: Elevated De Ritis Ratio ≥ 1.0 (Yes vs. No)]. Model goodness-of-fit was evaluated using Nagelkerke *R²* values, and Adjusted Odds Ratios (aOR) with corresponding 95% Confidence Intervals (95% CI) were reported. All statistical tests were two-tailed, with significance set at *p* < 0.05.

## Results

### Socio-demographic characteristics of the study population

A total of 174 school-aged children and adolescents across four endemic communities was enrolled in the study (Table 1). The cohort comprised 89 males (51.1%) and 85 females (48.9%). Based on pediatric age stratification, preschool children aged 3–4 years accounted for 9.2% (n = 16) of the cohort, early primary children aged 5–9 years represented the largest proportion (n = 88, 50.6%), older children aged 10–12 years comprised 19.0% (n = 33), and adolescents aged 13–16 years accounted for 21.3% (n = 37). By geographic distribution, half of the study population was recruited from Tato Battor (n = 87, 50.0%), followed by Kpatsa (n = 39, 22.4%), Gobu (n = 35, 20.1%), and Kwaese (n = 13, 7.5%).

### Infection intensity distribution by age categories

Parasitological infection status, egg density parameters, and baseline demographic comparisons are summarized in Table 2. Of the 174 enrolled participants, active *S. haematobium* infection (microscopic egg detection) was identified in 84 individuals (48.3%), while 90 individuals (51.7%) were confirmed uninfected. Sampling occurred 3 months following a district-wide PZQ MDA campaign, although individual treatment ingestion history was not formally recorded. Overall egg burden among infected participants ranged from 1.00 to 86.00 eggs/10 mL (median: 4.50, IQR: 1.00–9.25 eggs/10 mL). When classified according to WHO infection intensity criteria, most infected children exhibited light-intensity infections (< 50 eggs/10 mL; n = 78, 44.8% of total cohort; 92.9% of infected cases). Heavy-intensity infection (≥ 50 eggs/10 mL) was identified in 6 participants (3.4% of total cohort; 7.1% of infected cases). Age distributions did not differ significantly between the uninfected (median: 8.00 years, IQR: 5.00–12.0) and infected (median: 9.00 years, IQR: 7.00–12.0) cohorts (Mann-Whitney U = 3389, p = 0.237).

**Table 2.**
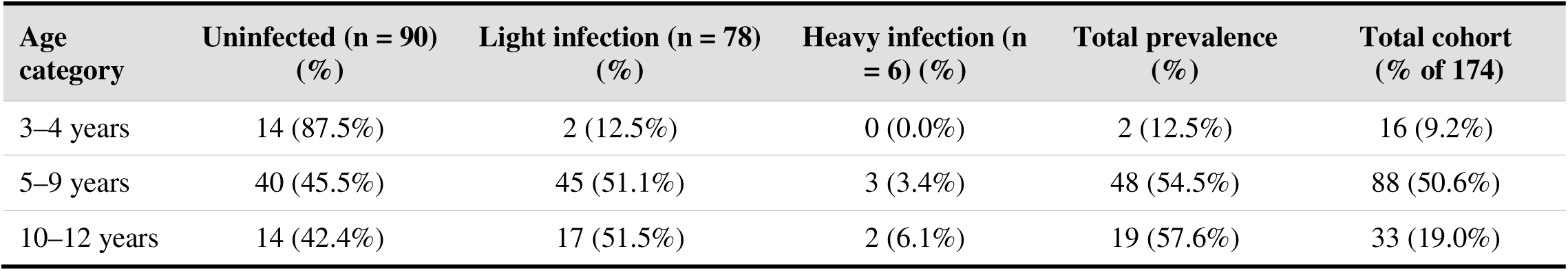
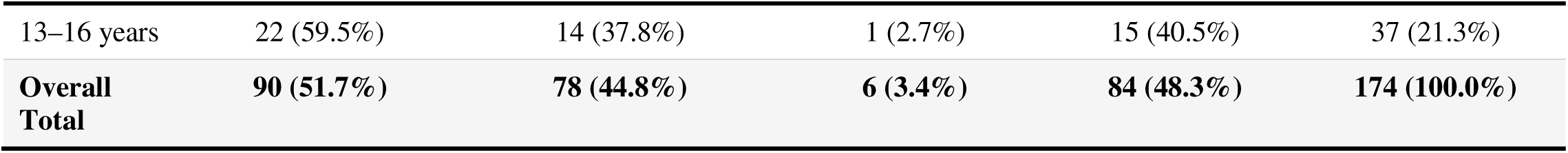
Distribution of *S. haematobium* infection status and WHO intensity categories across pediatric age groups (N = 174)

### Comparison of systemic biomarkers and electrolytes by infection status

Continuous laboratory biomarkers and serum electrolytes stratified by *S. haematobium* infection status are summarized in Table 3. Individual systemic hepatic enzyme levels showed no statistically significant differences in median values between uninfected and infected cohorts: alanine aminotransferase (ALT: 19.0 vs. 21.8 U/L; U = 3244, p = 0.107), aspartate aminotransferase (AST: 30.4 vs. 29.5 U/L; U = 3661, p = 0.720), alkaline phosphatase (ALP: 195 vs. 158 U/L; U = 3291, p = 0.141), gamma-glutamyl transferase (γ-GGT: 27.3 vs. 34.1 U/L; U = 3229, p = 0.097), and total bilirubin (9.30 vs. 10.3 µmol/L; U = 3245, p = 0.107).

**Table 3.** Continuous laboratory biomarkers and serum electrolytes stratified by *S. haematobium* microscopy status.

| Biomarker / Variable | Infection negative<br>(n = 90)<br>Median (Q1–Q3) | Infection positive<br>(n = 84)<br>Median (Q1–Q3) | Mann-Whitney U | p-value |
| --- | --- | --- | --- | --- |
| **Systemic hepatic biomarkers** |  |  |  |  |
| Alanine Aminotransferase (ALT; U/L) | 19.0 (13.9–27.2) | 21.8 (14.0–36.4) | 3244 | 0.107 |
| Aspartate Aminotransferase (AST; U/L) | 30.4 (21.1–36.8) | 29.5 (19.5–39.0) | 3661 | 0.720 |
| Alkaline Phosphatase (ALP; U/L) | 195 (106–307) | 158 (105–247) | 3291 | 0.141 |
| Gamma-Glutamyl Transferase (γ-GGT; U/L) | 27.3 (23.1–39.2) | 34.1 (23.3–51.4) | 3229 | 0.097 |
| Total Bilirubin (μmol/L) | 9.30 (6.65–13.9) | 10.3 (7.76–16.5) | 3245 | 0.107 |
| **Systemic renal biomarkers & electrolytes** |  |  |  |  |
| eGFR (mL/min/1.73m<sup>2</sup>) | 52.8 (44.3–69.5) | 54.9 (45.2–80.4) | 3507 | 0.412 |
| Serum Creatinine (μmol/L) | 82.0 (67.2–104) | 86.3 (63.3–102) | 3772 | 0.981 |
| Serum Urea (mmol/L) | 3.80 (3.23–4.50) | 3.87 (2.98–5.23) | 3643 | 0.680 |
| Serum Sodium (Na<sup>+</sup>; mmol/L) | 139 (138–141) | 139 (137–141) | 3651 | 0.890 |
| Serum Potassium (K<sup>+</sup>; mmol/L) | 4.10 (3.81–4.41) | 4.00 (3.84–4.40) | 3319 | 0.793 |
| Serum Chloride (Cl<sup>-</sup>; mmol/L) | 102 (101–106) | 104 (101–106) | 3319 | 0.165 |

*Note: Data presented as Median (Q1–Q3), where Q1 = 25th percentile and Q3 = 75th percentile. Group comparisons were performed using the non-parametric Mann-Whitney U test due to non-normality across continuous parameters ( $p < 0.05$ on Shapiro-Wilk test). Significance threshold was set at $p < 0.05$ . eGFR = Estimated Glomerular Filtration Rate; Na<sup>+</sup> = Sodium; K<sup>+</sup> = Potassium; Cl<sup>-</sup> = Chloride.*

Similarly, renal biomarkers and serum electrolytes maintained comparable median distributions across infection status: eGFR (52.8 vs. 54.9 mL/min/1.73 m²; U = 3507, p = 0.412), serum creatinine (82.0 vs. 86.3 µmol/L; U = 3772, p = 0.981), serum urea (3.80 vs. 3.87 mmol/L; U = 3643, p = 0.680), sodium (Na : 139 vs. 139 mmol/L; U = 3651, p = 0.890), potassium (K : 4.10 vs. 4.00 mmol/L; U = 3319, p = 0.793), and chloride (Cl : 102 vs. 104 mmol/L; U = 3319, p = 0.165).

### Prevalence of local urogenital indicators and systemic biomarker alterations

Evaluation of point-of-care urogenital dipstick indicators and systemic biomarker alteration frequencies stratified by *S. haematobium* infection status revealed marked localized differences between groups (Table 4).

**Table 4.** Baseline prevalence of point-of-care urinalysis indicators and composite systemic hepatorenal biomarker alterations stratified by *S. haematobium* infection status.

| Biomarker category / parameter | Overall (N=174) n (%) | Uninfected (n=90) n (%) | Infected (n=84) n (%) | Test Statistic ($\chi^2$) | p value |
| --- | --- | --- | --- | --- | --- |
| Local urogenital dipstick parameters |  |  |  |  |  |
| Microhematuria (Blood) |  |  |  |  |  |
| Negative | 116 (66.7%) | 80 (88.9%) | 36 (42.9%) | 41.40 | < .001* |
| Positive | 58 (33.3%) | 10 (11.1%) | 48 (57.1%) |  |  |
| Proteinuria |  |  |  |  |  |
| Negative | 118 (67.8%) | 70 (77.8%) | 48 (57.1%) | 8.48 | **0.004\*** |
| Positive | 56 (32.2%) | 20 (22.2%) | 36 (42.9%) |  |  |
| **Leukocyturia (Leukocytes)** |  |  |  |  |  |
| Negative | 147 (84.5%) | 86 (95.6%) | 61 (72.6%) | 17.40 | < **0.001\*** |
| Positive | 27 (15.5%) | 4 (4.4%) | 23 (27.4%) |  |  |
| **Systemic biomarker alterations** |  |  |  |  |  |
| **Hepatic biomarker alteration** |  |  |  |  |  |
| No (0) | 89 (51.1%) | 56 (62.2%) | 33 (39.3%) | 9.15 | **0.002\*** |
| Yes (1) | 85 (48.9%) | 34 (37.8%) | 51 (60.7%) |  |  |
| **Renal biomarker alteration** |  |  |  |  |  |
| No (0) | 134 (77.0%) | 72 (80.0%) | 62 (73.8%) | 0.94 | 0.332 |
| Yes (1) | 40 (23.0%) | 18 (20.0%) | 22 (26.2%) |  |  |
| **De Ritis Index** |  |  |  |  |  |
| Normal ratio (<1.0) | 130 (74.7%) | 72 (80.0%) | 58 (69.0%) | 2.76 | 0.097 |
| Elevated ratio (≥1.0) | 44 (25.3%) | 18 (20.0%) | 26 (31.0%) |  |  |

Note. Proportions present row percentages within infection status categories. Proportions compared using Pearson Chi-Square ( $\chi^2$ ) test or Fisher's Exact test. \* Bold p-value indicates statistical significance ( $p < .05$ ).

Assessment of individual urinalysis indicators showed that microhematuria was present in 33.3% (n = 58/174) of the overall cohort, with a significantly higher prevalence among infected participants (57.1%, n = 48/84) compared to uninfected participants (11.1%, n = 10/90; χ² = 41.40, p < 0.001). Proteinuria was observed in 32.2% (n = 56/174) of all individuals, demonstrating a significantly higher positivity rate in the infected group (42.9%, n = 36/84) than in the uninfected group (22.2%, n = 20/90; χ² = 8.48, p = 0.004). Similarly, leukocyturia was detected in 15.5% (n = 27/174) of the total study population, with infected participants displaying a significantly higher frequency (27.4%, n = 23/84) relative to the uninfected group (4.4%, n = 4/90; χ² = 17.40, p < 0.001).

Serum biomarker alterations were evaluated using predefined pediatric reference cutoffs (Methods). While univariate comparisons of individual serum biomarkers revealed subtle variations that did not achieve statistical significance independently (Table 3), composite panel analysis demonstrated a significant association for hepatic markers (Table 4). Specifically, composite hepatic biomarker alterations were identified in 48.9% (n = 85/174) of the cohort and were significantly associated with active infection (60.7%, n = 51/84 in infected participants vs. 37.8%, n = 34/90 in uninfected participants; χ² = 9.15, p = 0.002). Contrariwise, composite renal biomarker alterations (eGFR < 60 mL/min/1.73 m², serum creatinine > 100 µmol/L, or elevated urea) were present in 23.0% (n = 40/174) of participants, though prevalence did not differ significantly by infection status (26.2%, n = 22/84 vs. 20.0%, n = 18/90; χ² = 0.94, p = 0.332).

Additionally, 25.3% (n = 44/174) of the cohort presented with a De Ritis ratio (AST/ALT) ≥ 1.0, with infected participants showing a non-significant trend toward a higher prevalence of an elevated ratio compared to uninfected participants (31.0%, n = 26/84 vs. 20.0%, n = 18/90; χ² = 2.76, p = 0.097).

### Multivariable predictors of systemic biomarker alterations and De Ritis ratio

Multivariable binary logistic regression models were constructed to evaluate the independent associations of *S. haematobium* infection status, sex, and age with hepatic and renal biomarker alterations, and an elevated De Ritis ratio (≥ 1.0) (Table 5).

**Table 5.** Multivariable binary logistic regression models evaluating *S. haematobium* infection status and demographic predictors of composite systemic biomarker alterations and elevated De Ritis ratio (N = 174)

| Outcome Model / Predictor | Estimate (b) | SE | z | aOR | 95% CI | p value |
| --- | --- | --- | --- | --- | --- | --- |
| **Model 4A: Hepatic biomarker alterations (Nagelkerke $R^2 = 0.0726$)** |  |  |  |  |  |  |
| Infection (Pos vs Neg) | 0.931 | 0.313 | 2.975 | 2.537 | 1.374 – 4.690 | **0.003\*** |
| Gender (Male vs Female) | 0.222 | 0.312 | 0.712 | 1.249 | 0.677 – 2.300 | 0.476 |
| Age (years) | 0.001 | 0.047 | 0.032 | 1.001 | 0.914 – 1.100 | 0.975 |
| **Model 4B: Renal biomarker alterations (Nagelkerke $R^2 = 0.0228$)** |  |  |  |  |  |  |
| Infection (Pos vs Neg) | 0.334 | 0.364 | 0.918 | 1.397 | 0.684 – 2.854 | 0.359 |
| Gender (Male vs Female) | 0.459 | 0.367 | 1.251 | 1.583 | 0.771 – 3.251 | 0.211 |
| Age (years) | 0.018 | 0.055 | 0.334 | 1.018 | 0.915 – 1.134 | 0.738 |
| **Model 4C: Elevated De Ritis ratio ($\geq 1.0$) (Nagelkerke $R^2 = 0.0460$)** |  |  |  |  |  |  |
| Infection (Pos vs Neg) | -0.634 | 0.360 | -1.760 | 0.531 | 0.262 – 1.070 | 0.078 |

| **Outcome Model / Predictor** | **Estimate (b)** | **SE** | **z** | **aOR** | **95% CI** | **p value** |
| --- | --- | --- | --- | --- | --- | --- |
| Gender (Male vs Female) | -0.303 | 0.356 | -0.852 | 0.738 | 0.367 – 1.480 | 0.394 |
| Age (years) | 0.078 | 0.055 | 1.407 | 1.081 | 0.970 – 1.210 | 0.159 |

*Note. aOR = Adjusted Odds Ratio; CI = Confidence Interval; SE = Standard Error. Reference groups: Infection = Negative, Gender = Female. \* Bold p-value indicates significance ( $p < 0.05$ ).*

In Model 4A, active *S. haematobium* infection emerged as a significant independent predictor of hepatic biomarker alterations (Nagelkerke R² = 0.073). Infected participants exhibited more than two-fold increased odds of hepatic biomarker alterations compared to uninfected participants (aOR = 2.54, 95% CI: 1.37–4.69, p = 0.003). Demographic variables in this model, including male sex (aOR = 1.25, 95% CI: 0.68–2.30, p = 0.476) and age in years (aOR = 1.00, 95% CI: 0.91–1.10, p = 0.975), showed no significant association with hepatic biomarker alterations.

In contrast, Model 4B indicated that *S. haematobium* infection was not a statistically significant independent predictor of renal biomarker alterations (Nagelkerke R² = 0.023; aOR = 1.40, 95% CI: 0.68–2.85, p = 0.359). Neither male sex (aOR = 1.58, 95% CI: 0.77–3.25, p = 0.211) nor age (aOR = 1.02, 95% CI: 0.92–1.13, p = 0.738) independently influenced renal outcomes.

Finally, Model 4C examined predictors for an elevated De Ritis ratio (AST/ALT ≥ 1.0; Nagelkerke R² = 0.046). After adjusting for age and sex, active *S. haematobium* infection showed a non-significant trend (aOR = 0.53, 95% CI: 0.26–1.07, p = 0.078). Male sex (aOR = 0.74, 95% CI: 0.37–1.48, p = 0.394) and age (aOR = 1.08, 95% CI: 0.97–1.21, p = 0.159) similarly showed no significant independent associations.

## Discussion

The present study evaluated the relationships between *S. haematobium* infection status, local urogenital indicators, as well as individual and composite systemic hepatorenal biomarker alterations among school-aged children in the Sene West District of Ghana. Active *S. haematobium* infection was predominant among primary school children aged 5–9 years, who exhibited the highest prevalence (54.5%) and had the majority of heavy-intensity cases. Infection rates waned among older adolescents aged 13–16 years (40.5%). This age-intensity pattern reflects classic epidemiological dynamics in endemic West African settings, driven by intense water-contact exposure during recreational swimming and domestic chores (Grimes et al., 2015; Opare et al., 2025; van der Werf et al., 2003; Tetteh-Quarcoo et al., 2022). Also, elevated susceptibility in early childhood is exacerbated by immunologic naivety to schistosome antigens. This prevents effective natural clearance prior to repeated exposures or chemotherapeutic intervention (King, Dickman & Tisch, 2005).

Given that younger children bear the highest burden of active infection (Tetteh-Quarcoo et al., 2022), age might be expected to correlate directly with the severity of systemic biochemical indicators. However, in our multivariable logistic regression models that evaluated systemic biomarker alterations and elevated De Ritis ratios (AST/ALT ≥ 1.0), age did not serve as an independent predictor. This suggests that subclinical hepatorenal biomarker alterations in this pediatric cohort may be primarily driven by active infection status and localized inflammatory activity rather than chronological age.

*S. haematobium*-infected children exhibited significantly higher frequencies of microhematuria, proteinuria, and leukocyturia compared to uninfected children. These individual point-of-care urinalysis parameters are well-established diagnostic indicators of urogenital schistosomiasis, reflecting localized mucosal lesions and inflammatory responses within the lower urinary tract (Vere et al., 2025; Opare et al., 2025; van der Werf et al., 2003). Microhematuria is recognized as one of the most sensitive point-of-care indicators for active infection (WHO, 2022, Ochodo et al., 2015). However, in our study, microhematuria was detected in only 57.1% of infected participants. This lower-than-anticipated sensitivity can be contextualized by the timing of field sampling, which occurred in February 2026, three months after a regional mass drug administration (MDA) campaign with PZQ. Within this three-month post-treatment period, PZQ therapy effectively clears adult worm burdens and promotes mucosal healing, substantially reducing active urogenital vascular bleeding and microhematuria (Tetteh-Quarcoo et al., 2022; Koukounari et al., 2015). However, persistent egg deposition or early re-infection from ongoing water contact can yield positive egg counts before overt mucosal bleeding recurs (Grimes et al., 2015; Stete et al., 2012). Proteinuria serves as an additional indicator of localized protein leakage and residual cellular exudation secondary to persistent urothelial inflammation (Vere et al., 2025). Evaluating these individual dipstick parameters alongside egg counts provides a nuanced assessment of lower urinary tract inflammatory activity in post-MDA surveillance settings (Krauth et al., 2015).

Despite marked localized inflammatory activity detected on dipstick testing, individual systemic biomarkers remained stable across infected and uninfected children. For instance, individual serum transaminases (AST, ALT), renal panel markers (creatinine, urea, eGFR), and serum electrolytes displayed no statistically significant differences between infected children and their uninfected peers. These observations align closely with previous findings in Ghana (Ephraim et al., 2021; Darko et al., 2020). From a pathophysiological standpoint, pediatric *S. haematobium* infection targets the lower urogenital tract primarily and rarely provokes acute hepatic parenchymal necrosis or abrupt nephron loss in young children (King, Dickman & Tisch, 2005). Pronounced systemic elevations in serum urea and creatinine generally characterize chronic, late-stage adult disease (Barsoum, 2012). In adults, such elevations stem from decades of cumulative structural sequelae, including severe ureteral strictures, bilateral hydronephrosis, and bladder wall calcification (da Silva et al., 2013). In contrast, infected children in pediatric cohorts typically present with subclinical, early-stage inflammatory alterations rather than frank systemic organ failure (Darko et al., 2020).

In multivariable logistic regression modeling (Table 5), active *S. haematobium* infection predicted composite hepatic biomarker alterations (aOR = 2.54, 95% CI: 1.37–4.69, p = 0.003), whereas composite renal alterations showed no significant association. This divergence reflects pediatric schistosomiasis pathophysiology. Egg embolization through portosystemic anastomoses into the portal circulation provokes localized inflammatory responses in hepatic vascular beds (Gryseels et al., 2006). In the present study, stable renal biomarker levels (serum urea and creatinine) indicate that active *S. haematobium* infection in this pediatric cohort was not associated with detectable acute functional renal impairment or overt parenchymal injury. This finding aligns with pediatric surveys in endemic West African settings where lower urinary tract inflammatory changes predominate over functional renal impairment (Koukounari et al., 2007; Darko et al., 2020).

Evaluating point-of-care urinalysis alongside multivariable models demonstrates that active *S. haematobium* infection is associated with both localized urogenital inflammation and composite systemic biomarker alterations. Adjusting for age and sex, active infection remained an independent predictor of composite hepatic biomarker alterations (aOR = 2.54, 95% CI: 1.37– 4.69, p = 0.003). Microhematuria and proteinuria reflect immediate urothelial irritation from egg passage, whereas hepatic alterations indicate concurrent low-grade vascular responses in the portal system (Wilkins et al., 1979). Controlling for demographic covariates confirms that active infection independently influences these subclinical biomarker elevations.

These multivariable findings (Table 5) carry practical public health implications for pediatric schistosomiasis surveillance. Point-in-time microscopic egg counts measure immediate egg passage rather than cumulative tissue responses. Relying solely on single-specimen urine microscopy under-diagnoses low-intensity infections because daily egg excretion fluctuates substantially (Ochodo et al., 2015; Krauth et al., 2015) and drops as ova become trapped within granulomata (Gryseels et al., 2006) or after recent PZQ administration (Stete et al., 2012). Although multi-day consecutive urine sampling improves diagnostic sensitivity, field operations in school-based screening settings are typically constrained to single-specimen collection due to logistical and compliance barriers (WHO, 2022). In post-MDA endemic settings, combining point-of-care urinalysis with targeted liver panel profiling captures subclinical hepatic involvement (aOR = 2.54) that routine single-urine egg counts underrepresent (Gryseels et al., 2006; WHO, 2022). While *S. haematobium* primarily affects the urogenital tract, egg embolization into the portal circulation via portosystemic anastomoses induces low-grade granulomatous inflammation that rarely elevates individual transaminases beyond normal clinical limits (Gryseels et al., 2006). Consequently, composite biomarker profiling provides a superior readout of subclinical organ pathology when single-specimen microscopy misses low-density or sequestered infections (WHO, 2022). Importantly, subclinical hepatic elevations could also be influenced by concurrent, low-level *S. mansoni* co-infection, although preliminary screening in this endemic district revealed a very low *S. mansoni* prevalence (1.5%) in the pediatric population (Acheampong et al., unpublished data).

Several methodological limitations warrant consideration. First, the cross-sectional design precludes direct causal inferences; it cannot establish the precise temporal sequence connecting parasite exposure, mucosal urogenital inflammation and systemic biomarker alterations. Second, sampling occurred three months’ post-mass drug administration in communities with persistent freshwater reliance. Consequently, light egg counts may reflect early reinfection dynamics or low residual worm burdens rather than treatment-naive infection profiles. Third, single-specimen urine microscopy may have under-detected low-intensity infections due to diurnal egg shedding fluctuations. Future surveillance using circulating cathodic antigen assays (Ochodo et al., 2015) or molecular diagnostics (Weerakoon, Gordon & McManus) could resolve this diagnostic constraint. Fourth, organ involvement was evaluated strictly via circulating biochemical panels without complementary structural imaging, such as renal-bladder ultrasonography (Serra et al., 2026). Finally, while the sample size provided adequate statistical power for multivariable regression modeling (Table 5), larger prospective cohorts are needed. Multi-center studies will help validate these categorical biomarker thresholds across broader endemic settings.

### Conclusion

Active *S. haematobium* infection in school-aged children independently predicts composite hepatic biomarker elevations, even when individual biomarker medians stay well within normal clinical thresholds. Thus, reliance on individual biomarkers fail to reveal this hepatic response in endemic communities. Although, non-invasive urine dipsticks readily identify localized active urogenital inflammation through microhematuria and proteinuria, multivariable modeling confirms that infection drives threshold-level hepatic biomarker deviations regardless of age or sex. Pairing routine point-of-care urinalysis with targeted serum biomarker profiling could provide control programmes with a practical strategy to track subclinical organ involvement and improve post-MDA morbidity surveillance in low-resource settings.

## Declaration

### Ethics approval and consent to participate

The study protocol was reviewed and approved by the Committee for Human Research and Ethics (CHRE) at the University of Energy and Natural Resources, Sunyani, Ghana (CHRE/CA/025/022). All eligible children aged 3–16 years residing in the selected communities were invited to participate in the study. Enrollment proceeded upon obtaining written informed consent from parents or legal guardians. In addition, written assent was sought from children aged 10–16 years to affirm their voluntary agreement to participate. Informed consent forms were signed or thumb-printed (in the presence of an impartial witness for non-literate parents/guardians) prior to specimen collection.

### Consent for publication

Not applicable.

### Availability of data and materials

Available from the corresponding author upon reasonable request.

### Authors’ contributions

**Conceptualization:** DKO, AA, KBO; **Data curation:** DKO, CWN, EAB, KBO; **Formal analysis: :** DKO, AA, CWN, EAB, KBO; **Investigation:** DKO, AA, P-CK, PN, CWN, EAB, YST, KBO; **Methodology:** DKO, AA, P-CK, PN, CWN, EAB, YST, KBO; **Resources:** DKO, KBO; **Supervision:** KBO; **Visualization:** DKO, CRC, KBO, ; **Writing – original draft:** DKO, KBO; **Writing – review & editing:** DKO, AA, P-CK, PN, CWN, EAB, YST, CRC, KBO.

## Data Availability

All data produced in the present work are contained in the manuscript

## Acknowledgements

We thank the health directorate, community leaders, school administrators, teachers, and participating children in the Sene West District for their support and cooperation in this research work.

## Declaration of generative AI and AI-assisted technologies in the manuscript preparation process

During the preparation of this work the authors used *Gemini* to solely for editorial language refinement and manuscript formatting. After using this tool/service, the authors reviewed and edited the content as needed and take full responsibility for the content of the published article.

